# Regulatory *de novo* mutations underlying intellectual disability

**DOI:** 10.1101/2022.11.14.22279410

**Authors:** Matias G De Vas, Fanny Boulet, Shweta S Joshi, Myles G Garstang, Tahir N. Khan, Goutham Atla, David Parry, David Moore, Inês Cebola, Shuchen Zhang, Wei Cui, Anne K Lampe, Wayne W Lam, Genomics England Research Consortium, Jorge Ferrer, Madapura M Pradeepa, Santosh S Atanur

**Author notes:** Corresponding author: **Dr Santosh Atanur**, 5^th^ Floor ICTEM building, Hammersmith hospital campus, Imperial College London, London, W12 0NN, UK, **Dr Madapura M Pradeepa**, Blizard Institute, Barts and The London School of Medicine and Dentistry, Queen Mary University of London, London, E1 2AT, UK. Contributed equally (Matias G De Vas, Fanny Boulet).

## Abstract

The genetic aetiology of a major fraction of patients with intellectual disability (ID) remains unknown. *De novo* mutations (DNMs) in protein-coding genes explain up to 40% of cases, but the potential role of regulatory DNMs is still poorly understood. We sequenced 63 whole genomes from 21 ID probands and their unaffected parents (trio). Additionally, we analysed 30 previously sequenced genomes from exome-negative ID probands. We found that regulatory DNMs were selectively enriched in fetal brain-specific and human-gained enhancers. DNM-containing enhancers were associated with genes that show preferential expression in the pre-frontal cortex, have been previously implicated in ID or related disorders, and exhibit intolerance to loss of function mutations. Moreover, we found that highly interacting regulatory regions from intermediate progenitor cells of the developing human cortex were strongly enriched for ID DNMs. Furthermore, we identified recurrently mutated enhancer clusters that regulate genes involved in nervous system development (*CSMD1, OLFM1*, and *POU3F3)*. The majority of the DNMs from ID probands showed allele-specific enhancer activity when tested using luciferase assay. Using CRISPR-mediated mutation and editing of epigenomic marks, we show that regulatory elements harbouring DNMs indeed function as enhancers and DNMs at regulatory elements affect the expression of putative target genes. Our results, therefore, provide new evidence to indicate that DNMs in fetal brain-specific enhancers play an essential role in the aetiology of ID.

## Introduction

Intellectual disability (ID) is a neurodevelopmental disorder characterised by limitations in intellectual functioning and adaptive behaviour (1). The clinical presentation of ID is heterogeneous, often coexisting with congenital malformations or other neurodevelopmental disorders such as epilepsy and autism (1), and the worldwide prevalence is thought to be near 1% (2). In the past decade, next-generation DNA sequencing has identified a large set of protein-coding genes underlying ID that harbour pathogenic *de novo* protein-truncating mutations and copy number variants (1,3). Nevertheless, despite this recent progress, only up to 40% of the ID cases can be explained by the *de novo* mutations (DNMs) in the protein-coding regions of the genome (4). DNMs located in non-coding regions of the genome could therefore account for some cases in which no causal pathogenic coding mutation has been identified.

Previous studies have implicated noncoding mutations in long-range *cis*-regulatory elements, also known as transcriptional enhancers, in monogenic developmental disorders, including preaxial polydactyly (*SHH*) (5,6), Pierre Robin sequence (*SOX9*)(7), congenital heart disease (*TBX5*) (8) and pancreatic agenesis (*PTF1A*) (9). Systematic analysis of mutations in evolutionarily ultra-conserved non-coding genomic regions has estimated that around 1-3% of patients with developmental disorders but lacking pathogenic coding mutations could carry pathogenic non-coding DNMs in fetal brain *cis*-regulatory elements (10). Moreover, large-scale whole-genome sequencing of patients with autism spectrum disorder (ASD) has demonstrated that DNMs in conserved promoter regions contribute to ASD, while no significant association was found between enhancer mutations and ASD (11).

Despite these precedents, efforts to implicate enhancer mutations in human disease face numerous challenges. Importantly, it is currently not possible to readily discern functional enhancer mutations from non-functional or neutral variants based on sequence features. This can be partially addressed through experimental analysis of regulatory DNA mutations. Moreover, we still need a complete understanding of which regulatory regions and which subsequences within the regulatory regions are most likely to harbour disruptive mutations. In addition, one of the biggest challenges in interpreting mutations in regulatory regions is correctly associating regulatory elements with the potential target genes. Systematic identification of tissue-specific promoter-enhancer interaction maps would help identification of regulatory regions that are associated with disease-relevant genes.

ID is a severe early-onset neurodevelopmental phenotype; hence we hypothesised that ID could result from DNMs in enhancers that are specifically active during fetal brain development rather than the adult brain. Furthermore, more than half of the human enhancers have evolved recently; thus evolutionarily not conserved (12), and advanced human cognition has been attributed to fetal brain enhancers that are gained during evolutionary expansion and elaboration of the human cerebral cortex (13). Therefore, we have reasoned that the regulatory sequences critical for intellectual functions may show sequence constraints within human populations regardless of their evolutionary conservation.

In this study, we deployed whole-genome sequence analysis, integrative genomic and epigenomic studies, together with experimental functional validations to show that DNMs in patients with ID are selectively enriched in fetal brain enhancers. We further show that DNMs map to enhancers that interact with known ID genes, genes that are intolerant to mutations, and genes specifically expressed in the pre-frontal cortex. Furthermore, we identify three fetal brain-specific enhancer domains with recurrent DNMs and provide experimental evidence that candidate mutations alter enhancer activity in neuronal cells. Using dual luciferase assay, we show that the majority of enhancer DNMs have allele-specific activity. Furthermore, using CRISPR/Cas9 mediated editing, we show that enhancer DNM results in the downregulation of target gene expression. Our results provide a new level of evidence that supports the role of DNMs in neurodevelopmental enhancers in the aetiology of ID.

## Results

### Whole-genome sequencing and identification of *de novo* mutations

We performed whole-genome sequencing (WGS) of 63 individuals, including 21 probands with severe intellectual disability (ID) and their unaffected parents, at an average genome-wide depth of 37.6X **(Table S1)**. We identified, on average, 4.18 million genomic variants per individual that included 3.37 million single nucleotide variants (SNVs) and 0.81 million short indels **(Table S1)**. We focused our analysis on *de novo* mutations (DNMs), including *de novo* copy number variant (CNV), as it has been shown that DNMs contribute significantly to neurodevelopmental disorders (3,14,15). We identified 1,261 DNMs (de novo SNVs and Indels) in 21 trios. An average of 60 high-quality DNMs were identified per proband including 55.2 SNVs and 4.8 indels per proband **(Table S2)**, which was similar to the number of DNMs identified per proband in previous WGS studies on neurodevelopmental disorders (3,11,16). We identified three *de novo* CNVs in our ID probands (**Table S3**), which is approximately 0.14 *de novo* CNVs per proband. The number of *de novo* CNVs per proband was similar to the expected number of *de novo* CNVs per individual (17).

### Protein-coding *de novo* mutations and copy number variants

The role of protein-truncating mutations in ID is well established. Hence, we first looked at DNMs located in protein-coding regions of the genome. A total of 23 DNMs were located in protein-coding regions (an average of 1.1 DNMs per proband). Of the 23 coding mutations, 15 were non-synonymous coding mutations or protein-truncating mutations. In four ID probands (19% of all analysed probands), we identified various potentially pathogenic mutations in the genes *KAT6A, TUBA1A, KIF1A*, and *NRXN1*, all of them previously implicated in ID (1,3). The mutation in *KAT6A* resulted in a premature stop codon, while genes *TUBA1A* and *KIF1A* showed non-synonymous coding mutations, which have been reported as likely pathogenic and pathogenic, respectively in ClinVar (18) **(Table S4)**. One *de novo* CNV resulted in the partial deletion of *NRXN1*, a known ID gene. A family with two affected siblings was analysed for the presence of recessive variants. These findings are in agreement with previous reports on the pathogenic role of DNMs in the ID (15). All the coding DNMs were confirmed by sanger sequencing (data not shown).

### ID *de novo* mutations are preferentially located in fetal brain-specific enhancers

In our severe ID cohort, we did not identify pathogenic coding DNMs in 17 ID cases (∼81%), hence we decided to investigate potentially pathogenic mutations in disease-relevant enhancer regions. Our cohort size was relatively small, hence to increase the sample size for statistical analysis, we included 30 previously published severe ID samples in which no pathogenic protein-coding DNMs have been found using WGS (3), yielding a total of 47 exome-negative ID cases.

We hypothesised that DNMs in fetal brain-specific enhancers (FBSE) could perturb expression levels of genes that are important for brain development, leading in this way to ID. We, therefore, identified 27,420 fetal brain-specific enhancers using the data from the Roadmap Epigenomics project (19) (see Methods). The majority (76.52%) of these fetal brain-specific enhancers were found to be candidate cis-regulatory elements (ccREs) defined by Encyclopedia of DNA elements (ENCODE) (20), confirming the regulatory role of these enhancers. In addition, we analysed 8,996 human brain-gained enhancers that are active during cerebral corticogenesis (13).

A total of 83 DNMs were located within fetal brain-specific enhancers or human gain enhancers (HGE, an average of 1.77 DNMs per proband), which include 82 *de novo* SNVs and one *de novo* Indel (**Table S5**). A total of 52 DNMs were located within fetal brain-specific enhancers, 30 DNMs were in human gain enhancers, while one overlapped with both fetal brain enhancers as well as a human gain enhancer. First, we investigated whether in our ID cohort (n=47) DNMs were enriched in the enhancers that are specifically active in the fetal brain or the enhancers that are active in specific subsections of the adult brain. As a control dataset, we used DNMs identified in healthy individuals in the Genome of Netherlands (GoNL) (21) study. In our ID cohort, DNMs were significantly enriched in fetal brain-specific enhancers and human gain enhancers compared to adult brain-specific enhancers (*P*=9.12 ×10^−7^; **Fig. 1a; Table S6**). On the contrary, in healthy individuals (GoNL), DNMs were depleted in fetal brain-specific enhancers compared to adult brain-specific enhancers (*P*=0.008; **Fig. 1a; Table S6**). When we analysed two ID cohorts, the in-house cohort (n=17) and severe ID cohort (n=30), separately, DNMs from both cohorts showed enrichment in fetal brain-specific enhancers as compared to adult brain enhancers (*P*=0.005 and 0.00045 respectively, **Fig. 1b**). This suggests that the enrichment of ID DNMs in the fetal brain enhancer was consistent across multiple cohorts and results obtained using a combined dataset (n=47) were not biased due to one specific cohort. Our results were consistent with the expectation that mutations in enhancers active during fetal brain development contribute to the aetiology of ID, a severe early-onset neurodevelopmental phenotype, rather than mutations in enhancers active in the adult brain.

**Figure 1:**
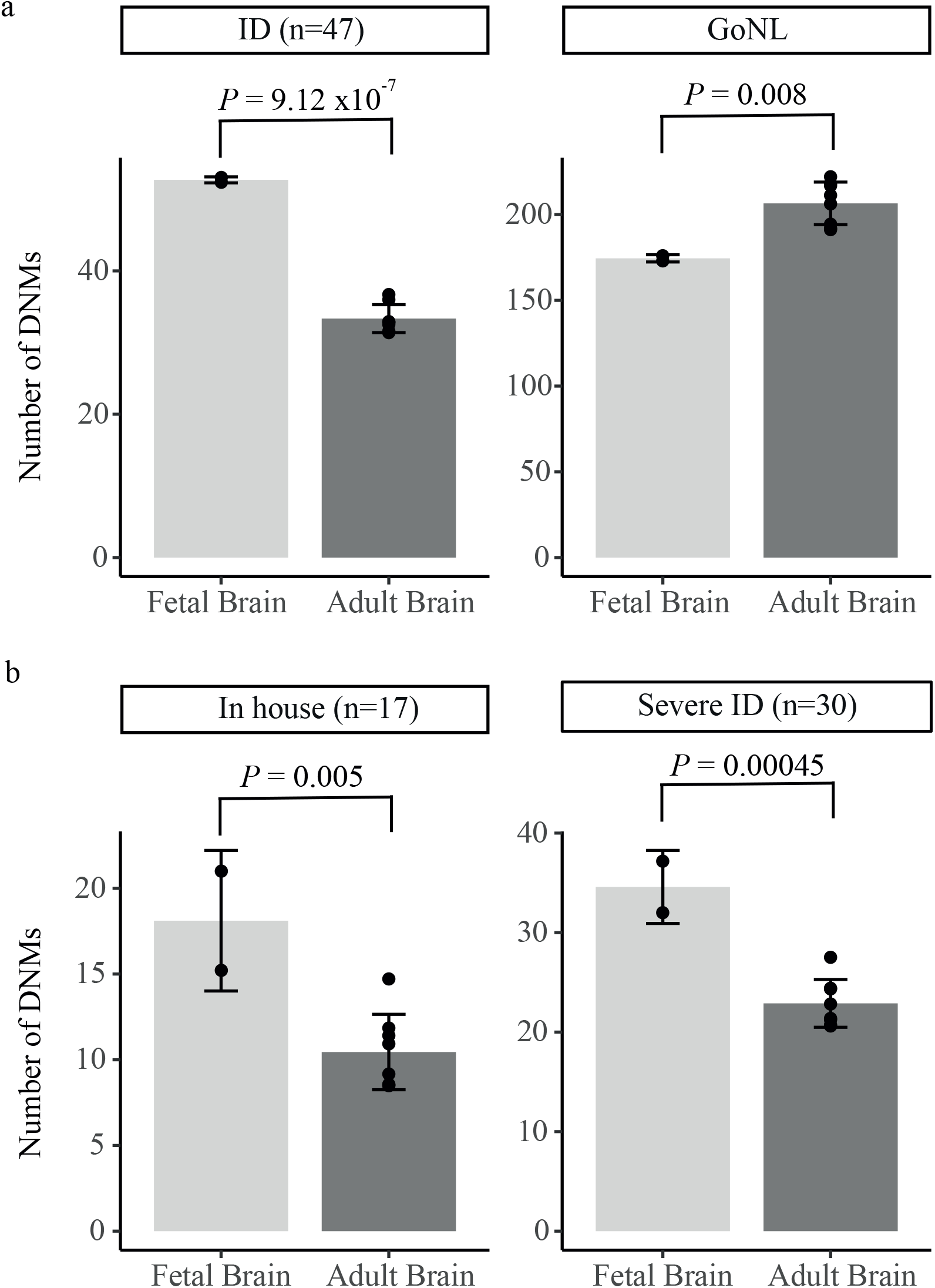
Enrichment of *de novo* mutations (DNMs) in fetal brain-specific enhancers. a) Enrichment of observed number of DNMs in fetal brain-specific enhancers and human gain enhancers as compared to adult brain sub-section specific enhancers in intellectual disability cohort (n=47) and healthy controls (GoNL). b) DNM enrichment in in-house (n=17) and severe ID cohort (n=30). The fetal brain enhancers (dark grey bars) represent fetal brain-specific and human-gained enhancers. The adult brain enhancers (light grey bar) represent adult brain sub-section specific enhancers, including angular gyrus, anterior caudate, cingulate gyrus, germinal matrix, hippocampus middle, inferior temporal lobe, dorsolateral prefrontal cortex, and substantia nigra. ID: Intellectual disability, GoNL: Genomes of the Netherland, In house: ID cohort sequenced in this study, Severe ID: Coding negative ID cohort from Gilissen et al, 2014 study.

### DNM-containing enhancers were associated with ID-relevant genes

We next investigated the hypothesis that DNMs are preferentially located in enhancers connected with genes that are plausible etiological mediators of ID. To identify potential target genes of fetal brain enhancers, we used the following datasets in sequential order: promoter capture Hi-C (PCHi-C) (22) from neuronal progenitor cells (NPC); correlation of H3K27ac ChIP-seq signal at promoters and enhancers across multiple tissues; and promoter-enhancer correlation using chromHMM segmentation data (19). The closest fetal brain expressed gene was assigned as a target gene for the 24% of the enhancers that remained unassigned after applying these approaches. For all approaches, we restricted our search space to topologically associated domains (TADs) defined in fetal brain tissue (23) as most enhancer-promoter interactions are restricted by TAD boundaries (24).

Next, we compiled a list of genes that have previously been implicated in ID or related neurodevelopmental disorders, using four gene sets: known ID genes (1,3), ID genes from Genomics England panel app (https://panelapp.genomicsengland.co.uk/), genes implicated in neurodevelopmental disorders in the Deciphering Developmental Disorder (DDD) project (14), and autism risk genes (SFARI genes) (25). The DNMs predominantly lead to dominant disorders; hence we selected dominant genes from the four gene lists for enrichment analysis. This resulted in a set of 617 dominant genes previously implicated in neurodevelopmental disorders. The genes associated with the DNM-containing fetal brain enhancers show significant enrichment for known neurodevelopmental disorder genes (17 genes, *P* = 5.4×10^−5^; **Table S7**). The most robust enrichment was observed for the DDD genes (15 genes, *P* = 1.05×10^−5^; **Table S7)**. Out of 47 coding negative ID patients in 17 patients, DNMs were observed in fetal brain-specific enhancers associated with known neurodevelopmental disorder genes.

We further observed that the target genes of DNM-containing enhancers were not only involved in nervous system development (*P* = 7.4×10^−4^; **Table S8**), but also predominantly expressed in the prefrontal cortex (*P* = 6.5 ×10^−3^; **Table S9**), a region of the brain that has been implicated in social and cognitive behaviour, personality expression, and decision-making.

The potential functional effect of enhancer mutations is expected to be mediated through the altered expression of target genes. Recently, it has been shown that most known severe haploinsufficient human disease genes are intolerant to loss of function (LoF) mutations (26). We compared the putative target genes of DNM-containing enhancers with the recently compiled list of genes that are intolerant to LoF mutations (pLI ≥ 0.9) (26). We found that a significantly higher proportion of enhancer DNM target genes were intolerant to LoF mutations than expected (*P* = 4.2×10^−5^; **Table S10**).

Taken together, our analysis shows that DNMs detected in severe ID patients are predominantly found in enhancers that regulate genes that are specifically expressed in the pre-frontal cortex, have been previously implicated in ID or related disorders, and exhibit intolerance to LoF mutations.

### ID *de novo* mutations in regulatory regions of human brain cell types

The human brain is a most complex tissue composed of multiple cell types and subtypes (27,28). We found that the genes associated with DNM-containing enhancers were predominantly expressed in the prefrontal cortex. The human cortex undergoes extensive expansion during development (29). The radical glia (RG), cortical stem cells give rise to intermediate progenitor cells (IPCs) and excitatory neurons (eN), which migrate to cortical plate (29,30), while interneurons (iN) migrate tangentially into the dorsal cortex (29,31). The developed human brain mainly consists of astrocytes, oligodendrocytes, microglia, neurons, and other cell types (27). It has been shown that the cell type-specific regulatory regions were enriched for the genome wide association studies (GWAS) risk variants for brain disorders and behavioural traits (27), hence we investigated regulatory regions in which specific human brain cell types are enriched for DNMs in the ID cohort. We obtained cell type-specific regulatory region annotations for developing human cortex (29) and developed human prefrontal cortex (27) from previous publications.

Cell type-specific open chromatin (ATAC-seq) data was available for radical glia, intermediate progenitor cells, excitatory neurons, and interneurons from developing cortex (29). We did not find enrichment for ID DNMs in open chromatin regions (ATAC-seq peaks) for any of the developing brain cell types (Table S11a). However, when the analysis was restricted to interacting open chromatin regions as defined by H3K4me3 mediated PLAC-seq, only interacting open chromatin regions of IPCs showed significant enrichment for ID DNMs compared to the GoNL DNMs (*P* = 5.18×10^−5^). Interestingly, this signal was driven by DNMs overlapping promoter regions rather than enhancers (Table S11b). The IPCs give rise to most neurons (32) hence DMNs in highly connected active promoters and enhancers from IPCs might have a profound impact on neurogenesis.

The enhancer regions of none of the four developed human brain cell types (astrocytes, oligodendrocytes, microglia, and neuronal cells), showed enrichment for ID DNMs compared to GoNL DNMs after multiple test corrections (Table 11c). Enhancers regions of only microglia (*P*= 0.0073) and neuronal cells (*P*=0.037) showed enrichment at a nominal p-value. On the contrary, all four developed human brain cell types showed significant enrichment for ID DNMs compared to GoNL DNMs in promoter regions after correcting for multiple tests. A total of 44 DNMs overlapped with the promoter regions of at least one of the four human brain cell types (Table 11d). Interestingly, the majority (70%) of the DNMs overlapped with the promoters that were active in all four cell types and 88.63% overlapped with promoters that were active in three or more cell types.

As enhancer regions of none of the human brain cell types showed significant enrichment for ID DNMs, we concentrated on DNMs overlapping enhancers from the bulk fetal brain for downstream analysis.

### Recurrently mutated enhancer clusters

In our cohort, we did not observe individual enhancers being recurrently mutated (containing two or more DNMs from unrelated probands). It has been shown that enhancers that regulate genes important for tissue-specific functions often cluster together (33). Therefore, we investigated whether clusters of fetal brain enhancers, i.e. sets of enhancers associated with the same gene, showed recurrent DNMs. We observed that the enhancer clusters associated with *CSMD1, OLFM1*, and *POU3F3* were recurrently mutated with two DNMs in each of their enhancer clusters **(Fig. 2)**. No DNMs were observed in the *CSMD1* and *OLFM1* enhancer clusters in healthy control individuals (GoNL), while four DNMs were observed in the *POU3F3* enhancer cluster in GoNL. The enrichment of DNMs in the *CSMD1* and *OLFM1* enhancer clusters was statically significant at a nominal p-value (Fisher’s exact test *P* = 0.02332). However, both clusters did not achieve a statistically significant p-value after multiple test corrections. The presence of three enhancer clusters with recurrent DNMs within the cohort of 47 ID probands was significantly higher than expected (permutation test *P* = 0.016). All three genes (*CSMD1, OLFM1*, and *POU3F3)* play a role in the nervous system development (34–36). Altogether, the known role of these genes in nervous system development and the presence of recurrent mutations in their enhancer clusters in the ID cohort suggest that these enhancer DNMs may contribute to ID.

**Figure 2:**
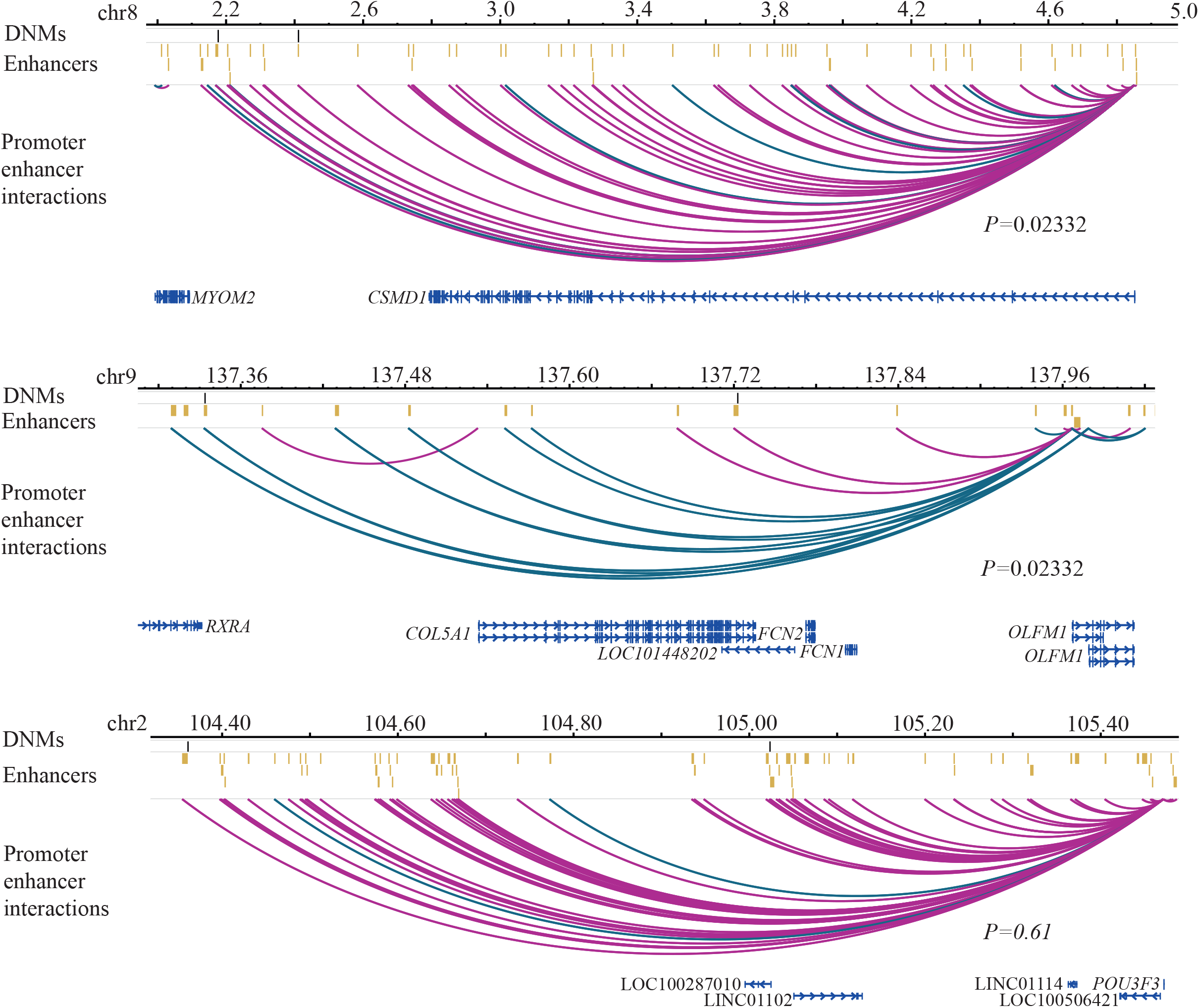
Enhancer clusters with recurrent DNMs in ID cohort. a) Recurrent DNMs in *the CSMD1* enhancer cluster. b) Recurrent DNMs in *the OLFM1* enhancer cluster. c) Recurrent DNMs in *the POU3F3* enhancer cluster. Black lines indicate DNMs, while yellow bars indicate enhancers. Pink arcs represent fetal brain-specific enhancer-promoter interactions, while green arcs represent human gain enhancer-promoter interactions. The scales are provided as million base pairs.

### DNMs in *CSMD1, OLFM1* and *POU3F3* enhancer clusters in a large ID cohort

To gain further insights into the enhancer clusters that showed more than one DNM in our cohort, we explored WGS data from large cohorts of neurodevelopmental disorders. Genomics England Limited (GEL) has sequenced 6,514 patients with intellectual disability. In GEL, no pathogenic coding variants were found in 3,169 ID patients for which WGS data of unaffected parents were available (trio WGS). We analysed DNMs from these 3,169 samples to find additional evidence supporting *CSMD1, OLFM1* and *POU3F3* enhancer DNMs. We found three individuals with *CSMD1* enhancer DNMs, five patients with DNMs in the *OLFM1* enhancer cluster, and 15 ID patients with DNMs in the *POU3F3* enhancer cluster. Next, we extracted human phenotype ontology (HPO) terms of the patients with DNMs in enhancers of these genes. Of three probands with *CSMD1* enhancer DNMs, two showed delayed speech and language development, delayed motor development, microcephaly, and seizures. All five probands with *OLFM1* enhancer DNMs showed delayed speech and language development, while three showed autistic behaviour and delayed motor development. Nine out of 15 probands with *POU3F3* enhancer DNMs showed autistic behaviour, global developmental delay and delayed speech and language development, while eight probands showed delayed motor development. Heterozygous mutations in *POU3F3* protein-coding regions have been recently implicated in ID (37,38). Phenotypes of probands with *POU3F3* enhancer mutations match the reported phenotypes of ID patients with coding *POU3F3* mutations (38). The phenotypic similarity between the patients harbouring DNMs in the enhancer of the same gene suggests they might be functional.

### Functional disruption of enhancer function by ID DNMs

Enhancers regulate gene expression through the binding of sequence-specific transcription factors (TFs) at specific recognition sites (39). DNMs could elicit phenotypic changes because they alter the sequence of putative TF binding sites or create putative TF binding sites (TFBS) that impact target gene expression. We used stringent criteria for TF motif prediction and motif disruption (see Methods). The software used to predict the effect of variants on TF motif (MotifbreakR) works only with single nucleotide variants (SNVs) not the Indels, hence we investigated only 82 *de novo* SNVs for their effect on TF motif disruption and excluded one *de novo* indel from this analysis. Of the 82 *de novo* SNVs that were located in fetal brain enhancers, 32 (39%) were predicted to alter putative TFBS affinity, either by destroying or creating TFBS **(Table S12a**). The fetal brain enhancer DNMs from ID probands frequently disturbed putative binding sites of TFs that were predominantly expressed in neuronal cells (*P* = 0.022; **Table S12b**). Our results suggest that the enhancer DNMs from ID probands were more likely to affect the binding sites of neuronal transcription factors and could influence the regulation of genes involved in nervous system development through this mechanism.

Of 17 in-house exome-negative probands, at least one DNM was predicted to alter TFBS affinity in 11 probands. To test the functional impact of regulatory mutations on enhancer activity, we randomly selected one DNM each from each of these 11 ID patients (**Table S12a**). Altogether we selected 11 enhancer DNMs **(Table S13)** and investigated their functional impact in luciferase reporter assays in the neuroblastoma cell line SH-SY5Y. Of the 11 enhancers containing DNMs, 10 showed significantly higher activity than the negative control (empty vector) in at least one allelic version (either wild type or mutant allele), indicating that they do indeed function as active enhancers in this neuronal cell line (**Fig. 3**). Amongst these 10 active fetal brain enhancers, nine showed allele-specific activity, with five showing loss of activity and four showing gain of activity of the DNMs **(Fig. 3)**. The *CSMD1* enhancer cluster had two DNMs (chr8:g.2177122C>T and chr8:g.2411360T>C) in two unrelated ID probands (Family 6 and Family 3, respectively). Both DNMs yielded a gain of enhancer activity compared to the wild-type allele (**Fig. 3**). By contrast, two DNMs in the *OLFM1* enhancer cluster (chr9:g.137722838T>G and chr9:g.137333926C>T) from two unrelated ID probands (Family 4 and Family 12, respectively) caused a loss of activity (**Fig. 3**). These results demonstrate that selected DNMs from ID patients in fetal brain enhancers alter predicted TF binding affinity and have a functional impact on enhancer activity assays.

**Figure 3:**
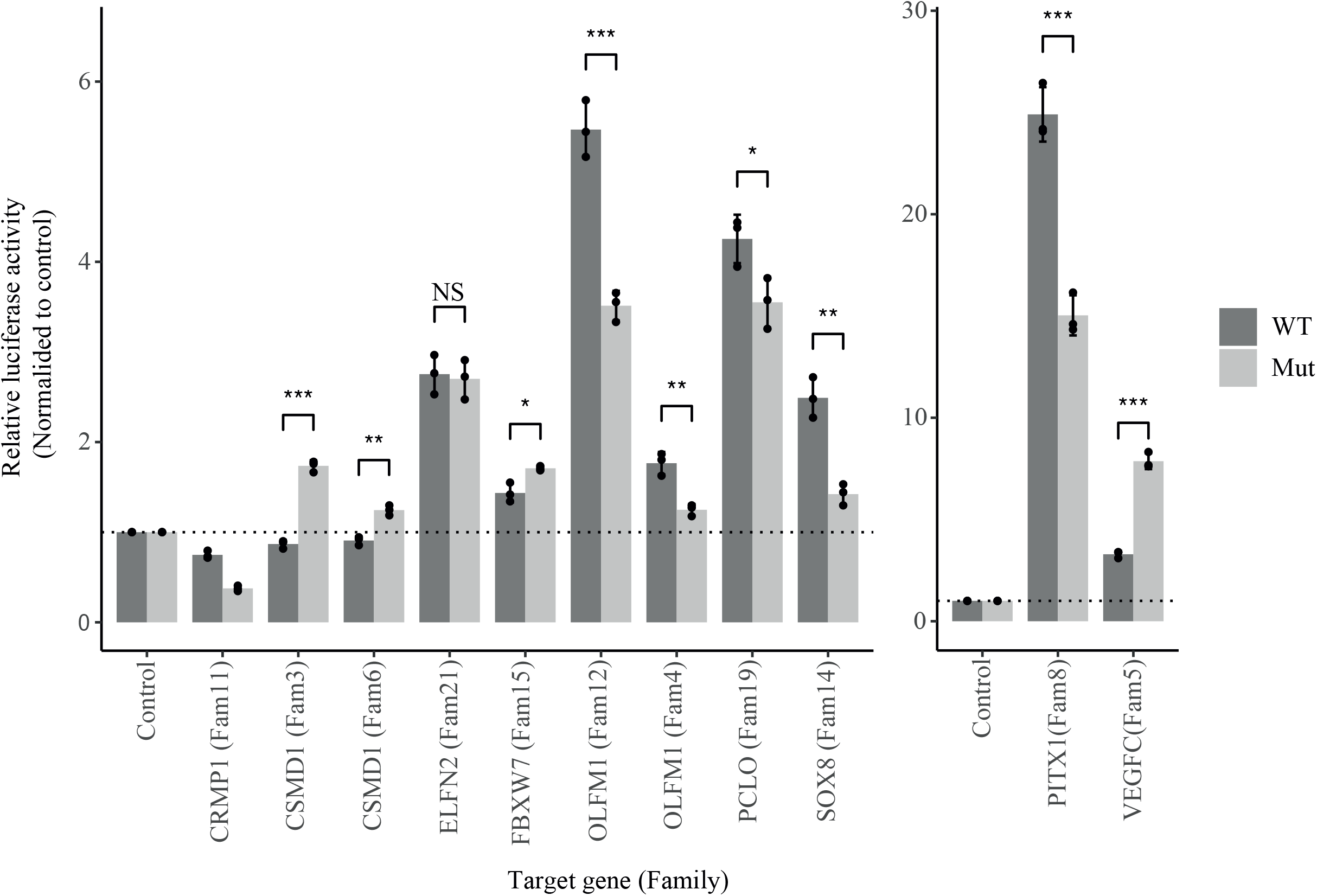
Effect of DNM on enhancer activity. Dual-luciferase reporter assay of wild type (reference) and the mutant (DNM) allele. The X-axis indicates the putative target genes of the enhancer, while the family IDs are shown in brackets. Y-axis indicates relative luciferase activity normalised to empty plasmid. The error bars indicate the standard error of means of three biological replicates. The enhancers associated with genes *PITX1* and *VEGFC* are plotted separately with different Y-axis scales because of the high activity of these enhancers. The significance level was calculated using a two-tailed t-test. *** Indicates p-value <=0.001, ** indicate p-value between 0.01 and 0.001 while * indicates p-value between 0.01 to 0.05.

### *SOX8* enhancer DNM leads to reduced expression of the *SOX8* gene

The luciferase reported assay is an episomal assay hence we randomly selected one DNM from the list of DNMs that showed allele-specific activity for investigation in genomic context using CRISPR. The luciferase reporter assays showed that the enhancer DNM (chr18:g.893142:C>A) from a family 14 proband results in a loss of enhancer activity (**Fig. 3**). The promoter capture HiC data from neuronal progenitor cells showed a strong interaction between DNM containing enhancer and the promoter region of *SOX8* (positive strand) and *LMF1* (negative strand) gene (**Fig. 4a**), suggesting that the *SOX8* and/or *LMF1* genes could be regulated by this enhancer in neuronal cells. The DNM containing enhancer was located approximately 139kb upstream of the *SOX8/LMF1* promoter (**Fig. 4a**). To investigate whether the putative enhancer of the *SOX8*/*LMF1* gene indeed regulates the expression of the target genes, we performed CRISPR interference (CRISPRi), by guideRNA mediated recruitment of dCas9 fused with the four copies of sin3 interacting domain (SID4x) in the neuronal progenitor cells (NPCs). We observed that CRISPRi of the *SOX8* enhancer led to downregulation of *SOX8* transcript levels in NPCs compared to non-target guideRNA controls (*P* = 0.034; **Fig. 4b**). This suggests that the DNM containing enhancer regulates the *SOX8* gene in neuronal cells.

**Figure 4:**
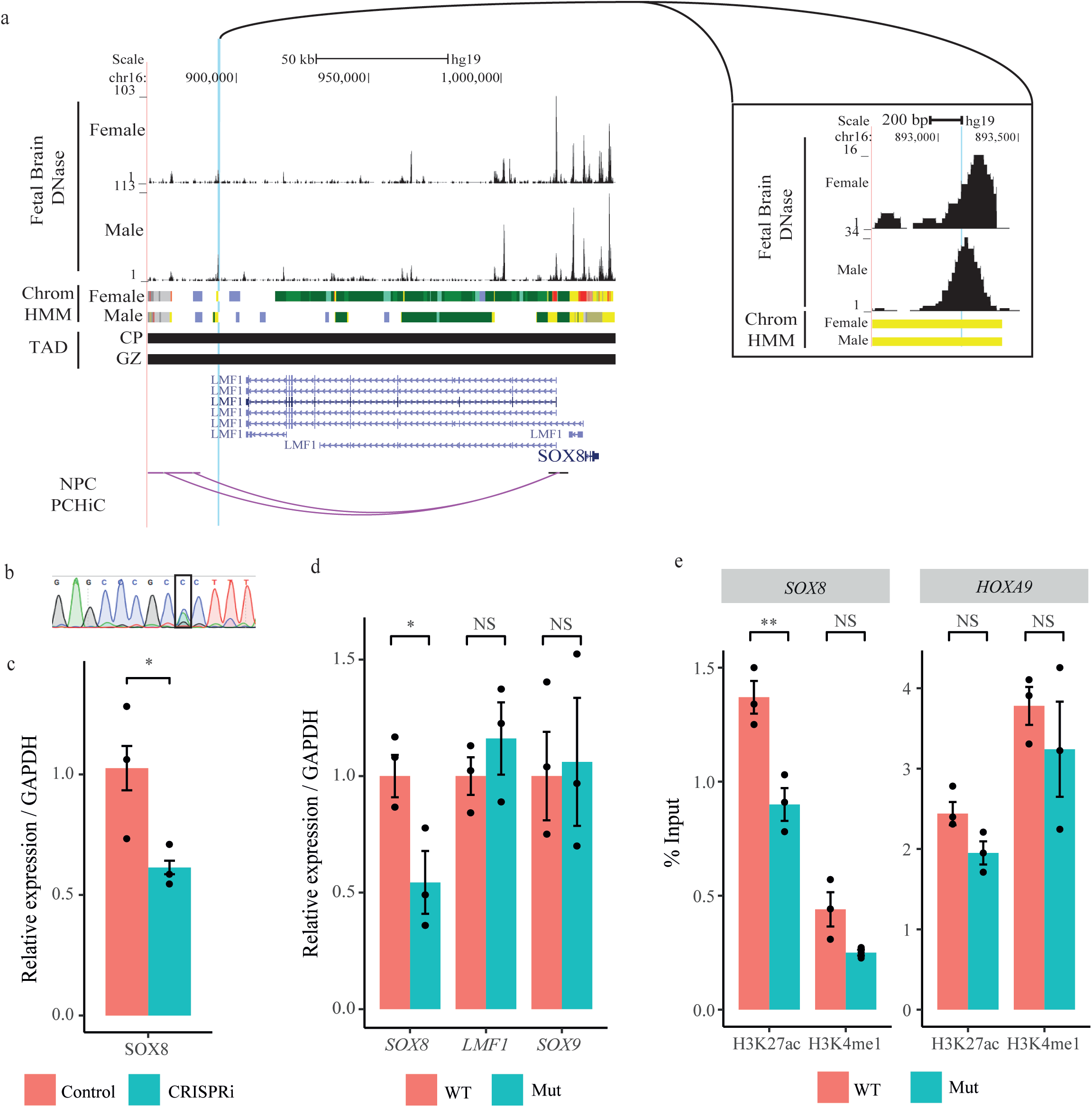
Characterisation of *SOX8* enhancer DNM using CRISPR. a) USCS tracks depicting male and female fetal brain DNase hypersensitivity peaks, ChromHMM tracks, fetal brain topologically associated domains (TAD), and enhancer-promoter interactions. Yellow bars in chromHMM tracks indicate enhancers. b) Relative expression levels of gene *SOX8* in neuronal progenitor cells (NPCs) with the CRISPRi of *SOX8* enhancer and non-target guide RNA controls, normalised to GAPDH c) Sanger sequencing trace file shows a heterozygous knock-in mutation in HEK293T cells using CRISPR/Cas9 mediated homology-directed repair. The black box highlights the location of the DNM. d) Relative expression levels of *SOX8, LMF1*, and *SOX9* in wild-type and mutant cell line normalised to GAPDH. e) Percentage of input (% input) of H3K27ac and H3K4me1 levels at *SOX8* enhancer and *HOXA9* promoter (control region) in wild-type and mutant cell line. The significance level was calculated using a two-tailed t-test. *** Indicates p-value <=0.001, ** indicate p-value between 0.01 and 0.001 while * indicates p-value between 0.01 to 0.05. WT: wild type, Mut: mutant.

The HEK293T cells show neuronal gene expression signature (40) and have been widely used for in vitro experiments to study neurodevelopment disorders (38). In addition, the DNM containing *SOX8* enhancer was active in HEK293T cells (**Fig. 4e**). To further investigate the direct impact of enhancer DNM on target gene expression in a genomic context, we knocked in enhancer DNM (chr18:g.893142:C>A) in the HEK293T cell line using CRISPR/Cas9 (**Fig. 4c**). In the heterozygous mutant clone, the *SOX8* gene showed a significant (*P* = 0.0301) reduction in expression levels, however, no difference was observed in expression levels of the *LFM1* gene (*P* = 0.8641; **Fig. 4d**), suggesting that the enhancer specifically regulates the *SOX8* gene but not the *LFM1* gene.

The presence of mono-methylation of histone H3 at lysine 4 (H3K4me1) and acetylation of histone H3 at lysine 27 (H3K27ac) is a strong indicator of active enhancers. The histone mark H3K4me1 can be observed at both active and inactive enhancers but the H3K27ac mark is observed only at active enhancers. Hence, we investigated H3K4me1 and H3K27ac levels at DNM containing *SOX8* enhancer. We observed a significant reduction in H3K27ac levels at the *SOX8* enhancer region (*P* = 0.0099) in the mutant clone as compared to the wild type. However, the level of H3K4me1 was not altered (*P* = 0.0674; **Fig. 4e**). The significant reduction in H3K27ac, which is associated with enhancer activity at DNM containing *SOX8* enhancer suggests a reduction in the enhancer activity upon DNM. At control loci (*HOXA9* promoter region) H3K27ac (*P* = 0.0751) and H3Kme1 (*P* = 0.444) levels were not altered in the mutant cells as compared to wild type. We, therefore, conclude that the enhancer harbouring DNM located 138,665bp downstream of the *SOX8* promoter regulates the expression of *SOX8* in neuronal cells.

Taken together, our analysis suggests that the *SOX8* enhancer mutation from family 14 ID proband indeed leads to reduced activity of the enhancers and, in turn, reduced expression of *the SOX8* gene. Further experimental analysis is required to establish the *SOX8* enhancer DNM as a cause of ID in this patient. However, our study provides strong evidence that *SOX8* enhancer DNM might be a causal variant in this patient.

## Discussion

Despite the current widespread use of whole-genome sequencing, the true burden of pathogenic mutations in enhancers is unknown. This is mainly due to an inability to predict the pathogenicity of enhancer mutations based on sequence features. Aggregation of a minority of pathogenic mutations with the majority of benign regulatory mutations nullifies any signal from pathogenic mutations in non-protein-coding genomic regions in disease cohorts. It is noteworthy that in protein-coding regions of the genome, only protein-truncating variants (PTV), but not other protein-coding mutations, show significant enrichment in neurodevelopmental disorders (11,41). The analysis of DNMs in selected monogenic phenotypes provides a powerful analysis instrument because it can focus on a relatively small number of mutations that are more likely to be pathogenic. In this study, we show that DNMs in a cohort of patients with ID exhibit a non-random genomic distribution that differs from DNMs observed in healthy individuals, with several features consistent with a pathogenic role of noncoding DNMs. DNMs from patients with ID were thus selectively enriched in fetal brain enhancers, enhancers associated with genes that are ID-relevant, intolerant to loss of function mutations, and genes specifically expressed in the pre-frontal cortex and disease-relevant transcription factor binding sites.

Identifying genes that are recurrently mutated across multiple individuals has been a major route to discovering novel disease genes (42). We identified recurrent mutations within three fetal brain enhancer clusters associated with genes involved in nervous system development (*CSMD1, OLFM1, POU3F3*) and found that this enrichment was significant relative to expectations. One of them (*POU3F3)* was recently shown to harbour pathogenic heterozygous mutations in patients with ID (38). The remaining two genes *CSMD1* and *OLFM1* show high pLI scores (pLI = 1), indicating that they are intolerant to loss of function (LoF) mutations and dosage sensitive. More than 72% of the genes that are intolerant to loss of function mutations have not been assigned to any human diseases despite the strong evidence of constraint (26). It has been speculated that heterozygous loss of function mutations in these genes might be embryonically lethal therefore loss of function mutations in these genes might not be observed in population (26). On the contrary, enhancers tend to be tissue, cell type, and developmental stage-specific hence effects of enhancer mutations might manifest only in the tissues or developmental stage at which the enhancer is active, leading to tissue-specific or developmental stage-specific disease phenotypes. We, therefore, hypothesise that DNMs in the tissue-specific enhancers of loss of function mutation intolerant genes might lead to disease even though the gene itself is not associated with any disease thus unravelling novel gene-disease association.

Our study is underpowered to perform any statistical analysis due to the smaller cohort size. Hence, we explored a large ID cohort from Genomics England to find evidence in support of our findings. Indeed, we observed multiple ID probands with DNMs in *CSMD1, OLFM1* and *POU3F3* enhancer clusters. Replicating our findings in a completely independent cohort provides strong support to our findings. Additionally, we performed extensive experimental validation of the fetal brain-specific enhancer DNMs observed in our cohort. We performed a dual-luciferase reporter assay for one enhancer DNM each from our coding negative probands that contain fetal brain-specific enhancer DNM. We found that most enhancer DNMs tested show a significant effect on the enhancer activity. The effect of DNMs was in both directions with an almost equal number of DNMs showing gain and loss of enhancer activity.

The *PITX1* enhancer with wild-type allele showed the highest activity amongst all the enhancers DNMs tested. A DNM from family 8 proband overlapping *PITX1* enhancer resulted in a significant loss of enhancer activity (*P*=0.0005). The enhancer was located upstream of the gene *PITX1*, within the same TAD as the gene. The deletions of regulatory regions upstream to *PITX1*, including family 8 DNM containing enhancer region, have been implicated in the Liebenberg syndrome (43–45). Haploinsufficiency of *PITX1* is the potential cause of Liebenberg syndrome (44). The significant reduction of *PITX1* enhancer activity due to DNM in our proband might lead to haploinsufficiency of the target gene (*PITX1*), which could cause disease phenotype in the family 8 proband.

Two DNMs from two probands were located on the *CSMD1* enhancer cluster. Both the DNMs located in *the CSMD1* cluster showed a significant effect on enhancer activity in the same direction, with the mutant allele showing a gain in enhancer activity (**Fig. 3**). The *CSMD1* gene is highly expressed in the central nervous system, particularly in the nerve growth cone (36). Common genomic variants in *CSMD1* are associated with schizophrenia and neuropsychological measures of general cognitive ability and memory function (39,40,41). Furthermore, *CSMD1* knockout mice show strong neuropsychological defects In our cohort, both ID probands with *CSMD1* enhancer DNMs showed developmental delay, and both were overgrown with high birth weights (above the 91st centile) and remained large throughout postnatal life. We identified three additional ID probands from Genomics England with DNM in the *CSMD1* enhancer cluster, that show similar phenotypes. The similarity in the phenotypes shown by both probands carrying *CSMD1* enhancer DNMs provides strong indications that the premature overexpression of *CSMD1* might cause ID in these patients.

Olfactomedin-1 (*OLFM1*), is a secreted glycoprotein expressed at high levels in the brain (50). It plays role in neural progenitor maintenance, cell death in the brain, and the optic nerve arborization (35). Translocation leading to *OLFM1* fusion protein has been implicated in Gilles de la Tourette syndrome, Obsessive-Compulsive Disorder and Attention deficit hyperactivity disorder (51). We identified two ID probands with DNMs in the *OLFM1* enhancer cluster in our cohort and five additional probands were found with *OLFM1* enhancer cluster DNMs in GEL. None of the individuals from the control cohort (GoNL) harboured DNM in the *OLFM1* enhancer cluster. This provides strong genetic evidence that DNMs in the OLFM1 enhancer cluster may play role in ID.

*POU3F3* is a well-known transcription factor involved in the development of the central nervous system (52). Recently, pathogenic DNMs in the *POU3F3* gene have been implicated in a novel neurodevelopmental disorder called Snijders Blok-Fisher syndrome (38). We have found two individuals with DNMs in the *POU3F3* enhancer cluster. Interestingly, 15 ID probands from GEL were also found to harbour DNM in the *POU3F3* enhancer cluster. The presence of 17 DNMs altogether in the *POU3F3* enhancer cluster and the phenotypic similarity between the patients harbouring coding and non-coding mutations suggests a potential role of *POU3F3* enhancer mutations in disease manifestation.

Luciferase assay is an episomal assay; hence we further investigated the effect of enhancer DNM on target gene expression in a genomic context. The *SOX8* is strongly expressed in embryonic and adult brain (53). We show that upon CRISPRi of putative *SOX8* enhancer transcript levels of *SOX8* were significantly reduced in NPCs suggesting the DNM containing enhancer regulates the *SOX8* gene in neuronal cells. In addition, we generated a heterozygous knock-in mutation at the putative *SOX8* enhancer in the HEK293T cell line. We show that the heterozygous variant at the putative *SOX8* enhancer region significantly reduces *SOX8* expression. This finding is particularly interesting as haploinsufficiency of *SOX8* has been implicated in ATR-16 syndrome characterised by alpha-thalassemia and intellectual disability (54,55). The *SOX8* DNM may, therefore, be a potential cause of ID in a proband from our cohort.

Our work has integrated whole-genome sequences, epigenomics, and functional analysis to examine the role of regulatory DNMs in ID. Despite the genetic heterogeneity of ID, which severely hampers efforts to unequivocal demonstrate a causal role for individual non-coding mutations, our results provide multiple lines of evidence to indicate that functional regulatory mutations in stage-specific brain enhancers contribute to the aetiology of ID. This work should prompt extensive genetic analyses and mutation-specific experimental modelling to elucidate the precise role of regulatory mutations in neurodevelopmental disorders.

## Materials and methods

### Selection criteria of intellectual disability patients for this study and ethical approval

The inclusion criteria for this study were that the affected individuals had a severe undiagnosed developmental or early-onset paediatric neurological disorder and that samples were available from both unaffected parents. Written consent was obtained from each patient family using a UK multicenter research ethics-approved research protocol (Scottish MREC 05/MRE00/74). The family IDs were provided randomly to each family, and they were not known to anyone outside the research group including clinicians.

### Sequencing and quality control

Whole-genome sequencing was performed on the Illumina X10 at Edinburgh Genomics. Genomic DNA (gDNA) samples were evaluated for quantity and quality using an AATI, Fragment Analyzer, and the DNF-487 Standard Sensitivity Genomic DNA Analysis Kit. Next-generation sequencing libraries were prepared using Illumina SeqLab-specific TruSeq Nano High Throughput library preparation kits in conjunction with the Hamilton MicroLab STAR and Clarity LIMS X Edition. The gDNA samples were normalised to the concentration and volume required for the Illumina TruSeq Nano library preparation kits, then sheared to a 450bp mean insert size using a Covaris LE220 focused-ultrasonicator. The inserts were ligated with blunt-ended, A-tailed, size selected, TruSeq adapters and enriched using 8 cycles of PCR amplification. The libraries were evaluated for mean peak size and quantity using the Caliper GX Touch with an HT DNA 1k/12K/HI SENS LabChip and HT DNA HI SENS Reagent Kit. The libraries were normalised to 5nM using the GX data, and the actual concentration was established using a Roche LightCycler 480 and a Kapa Illumina Library Quantification kit and Standards. The libraries were normalised, denatured, and pooled in eights for clustering and sequencing using a Hamilton MicroLab STAR with Genologics Clarity LIMS X Edition. Libraries were clustered onto HiSeqX Flow cell v2.5 on cBot2s, and the clustered flow cell was transferred to a HiSeqX for sequencing using a HiSeqX Ten Reagent kit v2.5.

### Alignment and variant calling

The de-multiplexing was performed using bcl2fastq (2.17.1.14), allowing 1 mismatch when assigning reads to barcodes. Adapters were trimmed during the de-multiplexing process. Raw reads were aligned to the human reference genome (build GRCh38) using the Burrows-Wheeler Aligner (BWA) mem (0.7.13) (56). The duplicated fragments were marked using samblaster (0.1.22) (57). The local indel realignment and base quality recalibration were performed using Genome Analysis Toolkit (GATK; 3.4-0-g7e26428) (58–60). For each genome, SNVs and indels were identified using GATK (3.4-0-g7e26428) HaplotypeCaller (61), creating a gvcf file. The gvcf files of all the individuals from the same family were merged and re-genotyped using GATK GenotypeGVCFs producing a single VCF file per family.

### Variant filtering

Variant Quality Score Recalibration pipeline from GATK (58–60) was used to filter out sequencing and data processing artefacts (potentially false positive SNV calls) from true SNV and indel calls. The first step was to create a Gaussian mixture model by looking at the distribution of annotation values of each input variant call set that match with the HapMap 3 sites and Omni 2.5M SNP chip array polymorphic sites, using GATK VariantRecalibrator. Then, VariantRecalibrator applies this adaptive error model to known and novel variants discovered in the call set of interest to evaluate the probability that each call is real. Next, variants were filtered using GATK ApplyRecalibration such that the final variant call set contains all the variants with a 0.99 or higher probability to be a true variant call.

### De novo mutations (DNM) calling and filtering

The de novo mutations (DNMs) were called using the GATK Genotype Refinement workflow. First, genotype posteriors were calculated using sample pedigree information, and the allele accounts from 1000 genome sequence data. Next, the posterior probabilities were calculated at each variant site for each trio sample. Genotypes with genotype quality (GQ) < 20 based on the posteriors are filtered out. All the sites at which the parents’ genotype and the child’s genotype with GQs >= 20 for each trio sample were annotated as the high confidence DNMs. we identified on average 1,527 DNMs in 21 probands (approximately 73 DNMs per proband) with GRCh38 assembly. Only high confident DNMs that were novel or had minor allele frequency less than 0.001 in 1000 genomes project were selected for further analysis. This resulted in the removal of 143 DNMs, resulting in a total of 1,384 DNMs (approximately 66 DNMs per proband).

Because most of the publicly available datasets, including epigenomic datasets, are mapped to human genome assembly version hg19, we lifted over all the DNM coordinates to hg19 using the liftover package. Liftover resulted in the loss of 123 DNMs as they could not be mapped back to hg19 resulting in a set of 1,261 DNMs (60 DNMs per proband). All the variant coordinates presented in this paper are from hg19 human genome assembly.

### DNM annotations

DNM annotations were performed using Annovar (62). To access DNM location with respect to genes, RefSeq, ENSEMBL, and USCS annotations were used. To determine allele frequencies, 1000 genome, dbSNP, Exac, and GnomAD databases were used. To determine the pathogenicity of coding DNMs, annotations were performed with CADD, DANN, EIGAN, FATHMM, and GERP++ pathogenicity prediction scores. In addition, we determined whether any coding DNM has been reported in the ClinVar database as a pathogenic mutation.

### Structural variant detection and filtering

To detect structural variants (SV), we used four complementary SV callers: BreakDancer v1.3.6 (63), Manta v1.5.0 (64), CNVnator v0.3.3 (65), and CNVkit v0.9.6 (66). The BreakDancer and Manta use discordantly paired-end and split reads to detect deletions, insertions, inversions, and translocations, while CNVnator and CNVkit detect copy number variations (deletions and duplications) based on read-depth information. The consensus SV calls were derived using MetaSV v0.4 (67). The MetaSV is the integrative SV caller, which merges SV calls from multiple orthogonal SV callers to detect SV with high accuracy. We selected SVs that were called by at least two independent SV callers out of four.

To detect *de-novo* SV, we used SV2 v1.4.1 (68). SV2 is a machine-learning algorithm for genotyping deletions and duplications from paired-end whole-genome sequencing data. In *de novo* mode, SV2 uses trio information to detect *de novo* SVs at high accuracy.

### Tissue-specific enhancer annotations

Roadmap Epigenomic Project (19) chromHMM segmentations across 127 tissues and cell types were used to define brain-specific enhancers. We selected all genic (intronic) and intergenic enhancers (“6_EnhG and 7_Enh) from a male (E081) and female fetal brain (E082). This was accomplished using genome-wide chromHMM chromatin state classification in rolling 200bp windows. All consecutive 200bp windows assigned as an enhancer in the fetal brain were merged to obtain enhancer boundaries. A score was assigned to each enhancer based on the total number of 200bp windows covered by each enhancer. Next, for each fetal brain enhancer, we counted the number of 200bp segments assigned as an enhancer in the remaining 125 tissues and cell types. This provided enhancer scores across 127 tissues and cell types for all fetal brain enhancers. To identify fetal brain-specific enhancers, Z scores were calculated for each fetal brain enhancer using the enhancer scores. Z scores were calculated independently for male and female fetal brain enhancers. Independent Z scores cutoffs were used for both male and female fetal brain enhancers such that approximately 35% of enhancers were selected. To define open accessible chromatin regions within brain-specific enhancers, we intersected enhancers with DNAse-seq data from Roadmap Epigenomic Project (19) from a male (E081) and female fetal brain (E082), respectively. Next, the male and female fetal brain-specific enhancers were merged to get a final set of 27,420 fetal brain-specific enhancers. We used a similar approach to identify enhancers that were specifically active in adult brain sub-sections, which include angular gyrus (E067), anterior caudate (E068), cingulate gyrus (E069), germinal matrix (E070), hippocampus middle (E071), inferior temporal lobe (E072), dorsolateral prefrontal cortex (E073) and substantia nigra (E074).

### Human gain enhancers

Human gain enhancers published previously by Reilly et al (13) were downloaded from Gene Expression Omnibus (GEO) using accession number GSE63649.

### *De novo* mutations from healthy individuals

We downloaded *de novo* mutations identified in the healthy individuals in genomes of the Netherland (GoNL) study (21) from the GoNL website.

### Fetal brain-specific genes

Roadmap Epigenomic Project (19) gene expression (RNA-seq) data from 57 tissues was used to identify fetal brain-specific genes. We used female fetal brain gene expression data, as RNA-seq data were available only for the female fetal brain. For each gene, Z scores were calculated using RPKM values across 57 tissues. The genes with a Z score greater than two were considered brain-specific.

### *De novo* mutation enrichment analysis

The size of enhancer regions differs widely between tissues. Furthermore, the mutability of the tissue-specific enhancer region differs significantly. The mutability of fetal brain-specific enhancers, human gain enhancers, and adult brain-specific enhancers was estimated using the previously defined framework for *de novo* mutations (69). The framework for the null mutation model is based on the tri-nucleotide context, where the second base is mutated. Using this framework, the probability of mutation for each enhancer was estimated based on the DNA sequence of the enhancer. The probability of mutation of all the enhancers within the enhancer set (fetal brain-specific enhancers, human gain enhancers, and adult brain sub-sections) was summed to estimate the probability of mutation for the entire enhancer set. To estimate the expected number of DNMs, the probability of mutation for each enhancer set was multiplied by the cohort size (n=47 for ID and n=258 for GoNL). The observed number of DNMs in all the enhancer sets was normalised to the expected number of mutations (mutability) of the fetal brain-specific enhancers. In short, to perform a valid comparison, we estimated the observed number of DNMs in all the enhancer sets used in this study to the same level of background mutability. The significance level was calculated using a two-tailed t-test.

### Enrichment of recurrently mutated enhancer clusters

The enhancer clusters were randomly shuffled 1000 times. We estimated the number of enhancer clusters with more than one mutation for each iteration. Then we counted the number of times more than or equal to two mutations were observed in three or more enhancer clusters. This number was then divided by 1000 to calculate P-value.

### DNM effect on transcription factor binding

The R Bioconductor package motifbreakR (70) was used to estimate the effect of DNM on transcription factor binding. The motifbreakR works with position probability matrices (PPM) for transcription factors (TF). MotifbreakR was run using three different TF databases: viz. homer, encodemotif, and hocomoco. To avoid false TF binding site predictions, either with reference allele or with alternate allele, a stringent threshold of 0.95 was used for motif prediction. DNMs that create or disturb a strong base (position weight >=0.95) of the TF motif, as predicted by motifbreakR, were selected for further analysis.

### Prediction of target genes of enhancers

Three different methods were used to predict the potential target genes of enhancers.

Chromosome conformation capture (Hi-C) comprehensively detects chromatin interactions in the nucleus; however, it is challenging to identify individual promoter-enhancer interactions using Hi-C due to the complexity of the data. In contrast, promoter capture Hi-C (PCHi-C) specifically identifies promoter-enhancer interactions as it uses sequence capture to enrich the interactions involving promoters of annotated genes (71). The significant interactions between promoters and enhancers identified using PCHi-C in neuronal progenitor cells (22) were used to assign target genes to the DNM-containing enhancers. The enhancers overlapped with the PCHi-C HindIII fragments. If an overlap was found between an enhancer and the PCHi-C HindIII fragment, the significantly interacting regions (PCHi-C HindIII fragments representing promoters of the genes) of the PCHi-C HindIII fragment were extracted to assign genes to the enhancers.

For an enhancer to interact with a promoter, both promoter and enhancer need to be active in specific cells at a specific stage. To identify promoter-enhancer interactions, all the active promoters in the fetal brain (as defined by chromHMM segmentation) were extracted. Promoter-enhancer interactions occur within topologically associated domains (TAD) hence, promoters located within the same TAD as that of a DNM-containing enhancer were used for analysis.

For each enhancer and promoter, H3K27ac counts were extracted from all tissues for which H3K27ac data was available in the Roadmap Epigenomic Project (19) ChIP-seq dataset. For the fetal brain, H3K27ac ChIP-seq data published by Reilly *et al* (13) was used because H3K27ac ChIP-seq data was not available in the Roadmap Epigenomics Project ChIP-seq dataset for the fetal brain. The Spearman rank correlation coefficient (Spearman’s rho) was calculated between each enhancer-promoter pair within the TAD using Scipy stats.spearmanr function from Python. The permutation test was performed to identify the significance of the correlation. The counts were randomly shuffled, independently for enhancers and promoters, 1000 times to calculate an adjusted *P*-value. The interactions with an adjusted *P* value less than 0.01 were considered a significant interaction between the enhancer and promoter.

Finally, if any enhancers remained unassigned to a gene using these approaches, they were assigned to the closest fetal brain-expressed genes within the TAD. A gene with an expression level more than or equal to 1 TPM in the Roadmap Epigenomics Project fetal brain RNA-seq data was considered to be expressed in the fetal brain.

### Gene enrichment analysis

To test if enhancer-associated genes were enriched for genes previously implicated in neurodevelopmental disorders, three different gene sets were used. 1) Intellectual disability (ID) gene list published in the review by Vissers *et al* (1) was downloaded from the Nature website 2) We compiled all the genes implicated in neurodevelopmental disorders in the Deciphering Developmental Disorder (DDD) project (14). 3) All the genes implicated in autism spectrum disorder were downloaded from the SFARI browser. The significance of enrichment was tested using a hypergeometric test in R.

Gene ontology enrichment and tissue enrichment analysis were performed using the web-based tool Enricher (http://amp.pharm.mssm.edu/Enrichr/).

The probability of loss of function intolerance (pLI) scores for each gene was downloaded from Exome Aggregation Consortium (ExAC) browser (http://exac.broadinstitute.org/). The significance of enrichment was tested using a hypergeometric test in R.

### Regulatory regions of developing human cortex and developed human pre-frontal cortex cell types

The developing human cortex cell type ATAC-seq and PLAC-seq data published in (29) was downloaded from the Neuroscience Multi-Omic Archive (NeMO Archive). The human pre-frontal cortex cell type enhancer and promoter annotations were downloaded from (27). DNMs were overlapped with regulatory regions using bedtools intersectBed (72). Significance of enrichment was calculated using fisher’s exact test in R.

### Cell culture

Neuroblastoma cell line (SH-SY5Y) was maintained in DMEM/F12 media (Gibco), 1% penicillin-streptomycin, 10 % fetal bovine serum, and 2 mM L-glutamine. The HEK293T cells were maintained in DMEM, 10% FCS, and 1% penicillin-streptomycin.

### Dual-luciferase enhancer assays

Enhancer and control regions (500-600 bp) were amplified from human genomic DNA from HEK293T cells using Q5 High-Fidelity Polymerase (NEB). Amplified fragments were cloned into pGL4.23 plasmid (Promega), which consists of a minimal promoter and the firefly luciferase reporter gene. These regions were mutagenised to introduce the *de novo* mutations of interest using the Q5 Site-Directed Mutagenesis kit (NEB) using non-overlapping primers. pGL4.23 plasmids containing putative enhancer DNA were sequence-verified and transfected, together with a Renilla luciferase-expressing vector (pRL-TK Promega) into SHSY-5Y cells using Lipofectamine 3000 (Invitrogen) following the manufacturer’s protocol. Firefly and Renilla luciferase activity was measured 24 hours after transfection using the Dual-Luciferase Reporter Assay System (Cat. number E1910, Promega) as per the manufacturer’s instructions. Primers used to amplify genomic DNA and for mutagenesis are provided in **Table S14**.

### Genome editing in HEK293T cells

To generate HEK293T cells carrying the mutation at the putative *SOX8* enhancer element, cells were co-transfected with gRNA (**Fig. S1**) expression plasmid (1□μg/ml) and the repair template with PAM mutation only (control) or repair template (1ul of 10 uM) with both PAM mutation and enhancer variant using Lipofectamine 3000 transfection reagent (Thermo Fisher) per the manufacturer’s instructions. After 48□h, successfully transfected cells were selected by puromycin treatment (2.5ug/ml) for 48 hours. The resulting puromycin-resistant cells were plated at 5,000□cells/10□cm2. After 1 week, colonies were picked and plated in duplicate at 1 colony/well of a 96-well plate. Genomic DNA was extracted from the colonies and sequenced by Sanger sequencing. Wild-type clones with PAM mutation only and heterozygous *SOX8* enhancer variant were expanded and frozen for later use. The *SOX8* enhancer sequencing primers are provided in **Table S15**.

### Genomic DNA extraction

Genomic DNA was extracted by a modified version of the salting-out method. Briefly, cells were lysed in Lysis Buffer (100 mM Tris-HCl pH 8.5; 5 mM EDTA; 200 mM NaCl; 0.2 % SDS) plus 4 U/mL of Proteinase K (Thermo Fisher) for at least 2 hours at 55ºC with agitation. Then, 0.4x volumes of 5 M NaCl were added to the mixture and centrifuged at max. speed for 10 min. DNA in the supernatant was precipitated with 1x volume of isopropanol. After centrifugation, the pellet was washed with 70% ethanol and air-dried for half an hour. DNA was resuspended in water and incubated for at least one hour at 37ºC with agitation.

### RNA isolation, cDNA synthesis, and RT-qPCR

RNA was extracted using the RNeasy Mini Kit (QIAGEN), and cDNA was produced with the First Strand cDNA Synthesis Kit (LunaScript^®^ RT SuperMix Kit, NEB). qPCR reactions were performed with SYBR Green Master Mix (Luna^®^ Universal qPCR Master Mix, NEB) and run on a CFX96 Real-Time PCR machine (Biorad). Relative gene expression values were calculated with the –ΔCt method, using GAPDH as a housekeeping gene for normalisation. Oligonucleotides used for qPCR are provided in **Table S16**.

### CRISPR interference (CRISPRi) with dCAS9-Sid4x

CRISPRi using dCAS9-Sid4x is performed as previously described in (73) with the following modifications. Oligos with guide RNA sequences (**Table S17**) were cloned into Addgene plasmid ID pSLQ1371 (kind gift by Stanley Qi) following the protocol previously described by (74).

For neural progenitor cells (NPCs) transfection, 7.5×10^5 NPCs were plated onto one well of a 6-well plate precoated with geltrex (Gibco, A1413302) in NPCs proliferation medium (49% Neurobasal medium (Gibco, A1647801), 49% Advanced DMEM/F12 (Gibco, 12634010) and 2% of Neural supplement (Gibco, A1647801). Approximately 24 hours after plating, 625ng of respective guide RNAs were diluted into 250μl opti-MEM (Gibco, 31985062) and 1875ng dCAS9-pSid4X. 10μl of TransIT-X2 reagent (Mirus Bio, MIR6004) was added to the above mix and thoroughly mixed and incubated at room temperature for 20 mins. 260μl of the final transfection mix was then added onto one well of a 6well plate containing 2.5ml media. The plate was rocked to mix and incubated for 24 hours. 48h after transfection, 0.5 μg/ml puromycin (Gibco, A1113803) was added to the media. Cells were harvested with accutase (Gibco, A1110501) after 24h of puromycin selection and taken for RNA isolation and qPCR.

For CRISPRi in NPC-derived neuronal cells, 10^5 NPCs were plated onto one well of a 24well plate precoated with 100μg/ml PLO (Sigma, P4957) and 5μg/ml Lamin (Sigma, L2020). Approximately 24 hours after plating, NPCs proliferation medium was replaced with 750-500ul Neuronal maturation medium (BrainPhys™ Neuronal Medium (Stemcell, 05790), NeuroCult™ SM1 Neuronal Supplement (Stemcell, 05711), N2 Supplement-A (Stemcell, 07152), 20ng/mL Human Recombinant BDNF (Stemcell, 78005), 20ng/mL Human Recombinant GDNF (Stemcell, 78058), 1mM Dibutyryl-cAMP (Sigma, D0627) and 200nM Ascorbic Acid (Stemcell, 072132)). 3days after the start of neuronal differentiation, 200ng of respective guide RNAs were diluted into 50μl opti-MEM and 600ng of dCAS9-pSid4X. 2μl of TransIT-X2 reagent was added to the above mix and thoroughly mixed and incubated at room temperature for 20 mins. 52μl of the final transfection mix was then added onto one well of a 24well plate containing 0.5ml Neuronal maturation medium. The plate was rocked to mix everything and incubated for 24 hours. 24h after transfection, 0.5 μg/ml puromycin was added to the media. Cells were harvested with accutase after 48h of antibiotic selection and taken for RNA isolation and qPCR.

### Statistical analysis

All luciferase experiments and gene quantification using qPCR were done in biological triplicates. The significance level was calculated using either a one-tailed or two-tailed t-test.

## Data Availability

The whole-genome sequence (WGS) data is not publicly available because it contains information that could compromise research participant privacy/consent. However, WGS data and variant calls that support the findings of this study are available on request from the corresponding author [S.S.A.].

## Declarations

### Ethics approval and consent to participate

Written consent was obtained from each patient family using a UK multicenter research ethics approved research protocol (Scottish MREC 05/MRE00/74).

## Acknowledgements

We thank the families of the affected children for their time and support for the research. We thank Prof Andrew Jackson for helpful discussions and for obtaining ethical approval for the study. We thank Mrs Sophie Shi for contributing to reagent generation. We also thank Dr Patrick Short and Dr Kaitlin Samocha both from Sanger Institute for providing a trinucleotide probability table and helpful discussion on a mutational model, respectively.

This research was made possible through access to the data and findings generated by the 100,000 Genomes Project. The 100,000 Genomes Project is managed by Genomics England Limited (a wholly owned company of the Department of Health and Social Care). The 100,000 Genomes Project is funded by the National Institute for Health Research and NHS England. The Wellcome Trust, Cancer Research UK and the Medical Research Council have also funded research infrastructure. The 100,000 Genomes Project uses data provided by patients and collected by the National Health Service as part of their care and support.

This research was supported by the National Institute for Health Research (NIHR) Imperial Biomedical Research Centre. This work was funded by grants from the Wellcome Trust Institute Strategic Support and National Institute for Health Research (NIHR) Imperial Biomedical Research Centre, Institute for Translational Medicine and Therapeutics (P70888) obtained by S.S.A. J.F.’s work was funded by grants from the Wellcome Trust (WT101033 to J.F.), Medical Research Council (MR/L02036X/1 to J.F.), European Research Council Advanced Grant (789055 to J.F.). M.M.P.’s lab is funded by the UKRI/MRC (MR/T000783/1), and Barts charity (MGU0475) grants. T.N.K. was partially supported by the Government of Pakistan under the PSDP project “Development of National University of Medical Sciences (NUMS), Rawalpindi”.

## Author contributions

Conceptualisation, S.S.A.; Acquiring funding, S.S.A.; Patient recruitment and sample collection, A.K.L, W.W.L. and D.R.F.; Bioinformatics analysis -Variant calling, S.S.J. and D.P.; Bioinformatics analysis: Enhancer-promoter interactions, G.A. and S.S.A.; Bioinformatics analysis: All other: S.S.A.; Experiments – Sanger sequencing: D.M.; Experiments – Luciferase: M.G.G., T.K., M.G.D., I.C., S.Z., and W.C.; Experiments – CRISPR: M.G.D., F.B. and M.M.P.; Writing – Original Draft: S.S.A.; Writing – Review & Editing, J. F., M.M.P., M.G.D., I.C., F.B., and M.G.G.; Supervision, D.R.F, J.F., M.M.P., and S.S.A.

## Abbreviations

ASD: Autism Spectrum Disorder
ATAC-seq: Assay for Transposase-Accessible Chromatin with high-throughput sequencing
BWA: Burrows-Wheeler Aligner
ccREs: candidate cis-Regulatory Elements
ChIP-seq: Chromatin ImmunoPrecipitation sequencing
CNV: Copy Number Variant
CRISPR: Clustered Regularly Interspaced Short Palindromic Repeats
CRISPRi: CRISPR interference
DDD: Deciphering Developmental Disorder
DNA: Deoxyribonucleic Acid
DNM: *De novo* Mutation
eN: excitatory Neurons
ENCODE: Encyclopedia of DNA Elements
ExAC: Exome Aggregation Consortium
FBSE: Fetal Brain-Specific Enhancers
GATK: Genome Analysis Toolkit
GEL: Genomics England Limited
GoNL: Genome of Netherlands
GQ: Genotype Quality
GWAS: Genome Aide Association Studies
H3K27ac: acetylation of histone H3 at lysine 27
H3K4me1: mono-methylation of histone H3 at lysine 4
H3K4me3: tri-methylation of histone H3 at lysine 4
HEK293T: Human Embryonic Kidney 293T
HGE: Human Gain Enhancers
Hi-C: Chromosome Conformation Capture
HPO: Human Phenotype Ontology
ID: Intellectual Disability
iN: interNeurons
Indels: Insertions and deletions
IPC: Intermediate Progenitor Cells
LoF: Loss of Function
PCHi-C: Promoter Capture Hi-C
PLAC-seq: Proximity ligation-assisted ChIP-Seq
pLI: probability of Loss of function Intolerance
PTV: Protein-Truncating Variants
RG: Radical Glia
SFARI: Simons Foundation Autism Research Initiative
SID4x: four copies of Sin3 Interacting Domain
SNV: Single Nucleotide Variant
TAD: Topologically Associated Domains
TF: Transcription Factors
TFBS: TF Binding Sites
TPM: Tags Per Million
VCF: Variant Call Format
WGS: Whole-Genome Sequencing

